# Genetic risk for Alzheimer’s disease influences neuropathology and cognition via multiple biological pathways

**DOI:** 10.1101/2020.07.16.20149658

**Authors:** Eilis Hannon, Gemma L Shireby, Keeley Brookes, Johannes Attems, Rebecca Sims, Nigel J Cairns, Seth Love, Alan J Thomas, Kevin Morgan, Paul T Francis, Jonathan Mill

## Abstract

Alzheimer’s disease is a highly heritable, common neurodegenerative disease characterised neuropathologically by the accumulation of β-amyloid plaques and tau-containing neurofibrillary tangles. In addition to the well-established risk associated with the *APOE* locus, there has been considerable success in identifying additional genetic variants associated with Alzheimer’s disease. Major challenges in understanding how genetic risk influences the development of Alzheimer’s disease are clinical and neuropathological heterogeneity, and the high level of accompanying comorbidities. We report a multimodal analysis integrating longitudinal clinical and cognitive assessment with neuropathological data collected as part of the Brains for Dementia Research (BDR) study to understand how genetic risk factors for Alzheimer’s disease influence the development of neuropathology and clinical performance. 693 donors in the BDR cohort with genetic data, semi-quantitative neuropathology measurements, cognitive assessments and established diagnostic criteria were included in this study. We tested the association of *APOE* genotype and Alzheimer’s disease polygenic risk score - a quantitative measure of genetic burden - with survival, four common neuropathological features in Alzheimer’s disease brains (neurofibrillary tangles, β-amyloid plaques, Lewy bodies and TDP-43 proteinopathy), clinical status (clinical dementia rating) and cognitive performance (Mini-Mental State Exam, Montreal Cognitive Assessment). The *APOE* ε4 allele was significantly associated with younger age of death in the BDR cohort. Our analyses of neuropathology highlighted two independent pathways from *APOE* ε4, one where β-amyloid accumulation mediates the development of tauopathy, and a second characterized by direct effects on tauopathy independent of β-amyloidosis. Although we also detected association between *APOE* ε4 and dementia status and cognitive performance, these were all mediated by tauopathy, highlighting that they are a consequence of the neuropathological changes. Analyses of polygenic risk score identified associations with tauopathy and β-amyloidosis, which appeared to have both shared and unique contributions, suggesting that different genetic variants associated with Alzheimer’s disease affect different features of neuropathology to different degrees. Taken together, our results provide insight into how genetic risk for Alzheimer’s disease influences both the clinical and pathological features of dementia, increasing our understanding about the interplay between *APOE* genotype and other genetic risk factors.

## Introduction

Alzheimer’s disease is a common neurodegenerative disease characterised clinically by progressive memory and cognitive decline leading to dementia and neuropathologically by β-amyloid plaques and tau-containing neurofibrillary tangles. The most frequent manifestation of Alzheimer’s disease is late onset Alzheimer’s disease (LOAD) where onset occurs after the age of 65. LOAD is highly heritable (Gatz *et al*., 2006) with the most established genetic risk factor being variants of the *APOE* gene. Relative to the most common genotype (ε3/ε3), the ε4 allele increases the risk of Alzheimer’s disease, with ε4 homozygosity associated with approximately 20-fold increase in risk (Farrer *et al*., 1997). In contrast, the ε2 allele of APOE has strong protective effects (Reiman *et al*., 2020). Genome-wide association studies (GWAS) in large sample cohorts (Lambert *et al*., 2013; Marioni *et al*., 2018; Jansen *et al*., 2019; Kunkle *et al*., 2019) have identified additional variants in more than 40 regions of the genome which individually confer subtler effects on risk, but cumulatively account for a large proportion of genetic risk. To index an individuals’ genetic risk profile, disease-associated variants - typically including those below genome-wide significance - can be combined into a ‘polygenic risk score’ (PRS). PRSs quantify the number of genetic risk variants an individual has, weighted by their effect size, and have been shown to improve prediction models of Alzheimer’s disease (Escott-Price *et al*., 2015; Cruchaga *et al*., 2018; Escott-Price *et al*., 2019). Of note, the Alzheimer’s disease PRS has greatest predictive power where disease status has been defined by standardized neuropathological assessment (Escott-Price *et al*., 2017), and is most elevated in sporadic early-onset cases (Cruchaga *et al*., 2018).

In addition to genetic prediction, PRSs provide a powerful mechanism to investigate how genetic risk mediates the development of symptoms, and can potentially be used to disentangle the primary causal features from the secondary consequences of disease. As well as being associated with dementia status, the Alzheimer’s disease PRS has been shown to correlate with mild cognitive impairment (Adams *et al*., 2015; Chaudhury *et al*., 2019), cognitive decline (Mormino *et al*., 2016; Marioni *et al*., 2017; Felsky *et al*., 2018), memory impairments (Mormino *et al*., 2016; Marioni *et al*., 2017), cortical thickness (Sabuncu *et al*., 2012; Corlier *et al*., 2018), hippocampal volume (Lupton *et al*., 2016; Mormino *et al*., 2016), cerebrospinal biomarkers (Martiskainen *et al*., 2015; Louwersheimer *et al*., 2016; Desikan *et al*., 2017), and neuropathology (Desikan *et al*., 2017; Felsky *et al*., 2018; Tasaki *et al*., 2018). The breadth of associations highlights the complexity of understanding the pathways from genetic risk to symptomatic disease. Furthermore, many of these analyses have included the *APOE* locus within the PRS, meaning their results may reflect *APOE-*specific effects rather than the consequences of a broader polygenic risk burden. To truly understand how multiple genetic risk factors combine to influence the interplay of the clinical, cognitive and neuropathological characteristics of Alzheimer’s disease, we need large, longitudinal cohorts with post-mortem tissue that can align genetics, clinical data and standardized neuropathological assessments.

A major challenge in understanding how genetic risk influences the development of Alzheimer’s disease relates to clinical and neuropathological heterogeneity, and the high level of accompanying comorbidities associated with a diagnosis of Alzheimer’s disease. The presence of the neuropathological hallmarks of Alzheimer’s disease can only be confirmed following post-mortem brain examination. Standardized sampling and staining methods, along with the introduction of a number of semi-quantitative classification schemes, each focused on a single neuropathological feature (Thal *et al*., 2002; Braak *et al*., 2003; Braak *et al*., 2006), promote consistency making it easier to harmonise data across brain banks and ultimately the reproducibility of findings across studies. It is now recognised that sporadic dementia in older people is predominantly due to multiple pathologies (Robinson *et al*., 2018). The most frequent comorbidity is Lewy body pathology affecting up to 50% of sporadic Alzheimer’s disease cases (Toledo *et al*., 2013). Another common comorbidity is the presence of inclusion bodies containing aggregates of transactive response DNA-binding protein 43 (TDP-43), particularly in the oldest old (Amador-Ortiz *et al*., 2007; Uryu *et al*., 2008; James *et al*., 2016). As well as influencing cognitive impairment in non-Alzheimer’s disease cases (Nag *et al*., 2017), these comorbidities contribute to the cognitive decline observed in Alzheimer’s disease cases beyond that associated with β-amyloid and neurofibrillary tangle pathology (Wilson *et al*., 2013) (Nelson *et al*., 2019), hence it is important to consider multiple neuropathological features simultaneously, to understand the processes that underlie cognitive performance in old age.

The paucity of comprehensive neuropathological data in large sample cohorts has limited previous genetic studies of Alzheimer’s disease-associated neuropathology. To address this gap, the Brains for Dementia Research (BDR) cohort was established in 2007 recruiting both dementia patients and unaffected controls over the age of 65 to partake in routine longitudinal assessments collecting cognitive, clinical, lifestyle and psychometric data, prior to post-mortem brain donation (Francis *et al*., 2018). The inclusion of standardized semi-quantitative data for a range of neuropathological features facilitates analyses into the specificity of genetic risk factors for the different abnormalities, and an assessment of their clinical contributions. In this study we report the first multimodal analysis of the BDR cohort, integrating longitudinal clinical and cognitive assessment with neuropathological data to explore how known genetic risk factors for Alzheimer’s disease influence the development of different aspects of neuropathology and cognitive performance in old age. We focus on four common neuropathological features observed in Alzheimer’s disease brain tissue: neurofibrillary tangles, β-amyloid plaques, Lewy bodies and TDP-43 proteinopathy. The results of this study provide insights into the neurobiological pathways to cognitive decline by refining our understanding of the complex interplay of genetic risk, clinical presentation and neuropathological burden.

## Materials and Methods

### Brains for Dementia Research (BDR) cohort description

BDR was established in 2007 and consists of a network of six dementia research centres in England and Wales (King’s College London, Bristol, Manchester, Oxford, Cardiff and Newcastle Universities) and the associated university brain banks handling the donations (Cardiff brain donations were banked in London). Participants over the age of 65 (including those with and without dementia) underwent a series of longitudinal cognitive and psychometric assessments, and registered for brain donation. An extensive description of the recruitment strategy, demographics, assessment protocols and neuropathic assessment procedures can be found in (Francis *et al*., 2018).

### Longitudinal cognitive and clinical assessments

All assessments were conducted by a trained psychologist or research nurse. Baseline assessments were conducted face-to-face (in the participant’s place of residence or a BDR centre), follow-up assessments were usually face-to-face but telephone interviews were also used for some healthy control participants. Follow-up interviews were annual for participants with cognitive impairment, and every 1 to 5 years (depending on age) for cognitively healthy participants. Clinical assessment was performed using the Clinical Dementia Rating (CDR) (Morris, 1993). Cognitive assessment measures relevant to this study included the Mini-Mental State Examination (MMSE) (Folstein *et al*., 1975) and Montreal Cognitive Assessment (MoCA) (Nasreddine *et al*., 2005).

### Post-mortem neuropathological assessment

After removal, the brain was examined macroscopically and digitally recorded. After slicing, the brain was comprehensively sampled according to the BDR protocol by experienced neuropathologists in each of the five network brain banks. This protocol, arrived at by consensus across the BDR network and based on the BrainNet Europe initiative (Bell *et al*., 2008), was used to generate a description of the regional pathology within the brain together with standardized scoring. In this study we considered five variables representing four neuropathological features: i) Braak tangle stage which captures the progression of neurofibrillary tangle pathology (Braak and Braak, 1991; Braak *et al*., 2006), ii) Thal β-amyloid phase which captures the regional distribution of plaques (Thal *et al*., 2002), iii) Consortium to Establish a Registry for Alzheimer’s Disease (CERAD) stage which profiles neuritic plaque density (Mirra *et al*., 1991; Montine *et al*., 2012), iv) Braak Lewy body stage (Braak *et al*., 2003) and v) TDP-43 status (a binary indicator of the absence/presence of TDP-43 inclusions, as assessed by immunohistochemistry of the amygdala and the hippocampus and adjacent temporal cortex for phosphorylated TDP-43). All variables apart from TDP-43 were analysed as continuous variables, using their semi-quantitative nature to capture dose-dependent relationships of increasing neuropathological burden.

### Genetic data

DNA extraction was performed using a standard phenol chloroform method on 100 mg of brain tissue. DNA quality was assessed using the Agilent 2200 TapeStation DNA integrity number and quantified using Nanodrop 3300 spectrometry. Genotyping was performed on the NeuroChip array which is a custom Illumina genotyping array with an extensive genome-wide backbone (*n* = 306,670 variants) and custom content covering 179,467 variants specific to neurological diseases (Blauwendraat *et al*., 2017). Genotype calling was performed using GenomeStudio (v2.0, Illumina) and quality control (QC) was completed using PLINK1.9 (Chang *et al*., 2015). Individuals were excluded if either 1) they had > 5% missing data, 2) their genotype predicted sex using X chromosome homozygosity was discordant with their reported sex (excluding females with an F value > 0.2 and males with and F value < 0.8), 3) they had excess heterozygosity (>3 SD from the mean), 4) they were related to another individual in the sample (pi hat > 0.2), where one individual from each pair of related samples was excluded considering data quality and phenotype, or 5) they were classed as non-European, determined by merging the BDR genotypes with data from HapMap Phase 3 (http://www.sanger.ac.uk/resources/downloads/human/hapmap3.html), linkage disequilibrium (LD) pruning the overlapping SNPs such that no pair of SNPs within 1500 bp had r^2^ > 0.20 and visually inspecting the first two genetic principal components along with the known ethnicities of the HapMap sample to define European samples (**Supplementary Fig. 1**). Prior to imputation SNPs with high levels of missing data (>5%), Hardy-Weinberg equilibrium P < 0.001 or minor allele frequency <1% were excluded. The genetic data were then recoded as vcf files before uploading to the Michigan Imputation Server (Das *et al*., 2016) (https://imputationserver.sph.umich.edu/index.html#!) which uses Eagle2 (Loh *et al*., 2016) to phase haplotypes, and Minimac4 (https://genome.sph.umich.edu/wiki/Minimac4) with the most recent 1000 Genomes reference panel (phase 3, version 5). Imputed genotypes were then filtered with PLINK2.0alpha, excluding SNPs with an R^2^ INFO score < 0.5 and recoded as binary PLINK format. Proceeding with PLINK1.9, samples with >5% missing values, and SNPs with >2 alleles, >5% missing values, Hardy-Weinberg equilibrium P < 0.001, or a minor allele frequency of <5% were excluded. The final quality controlled imputed set of genotypes contained 6,607,832 variants.

### Polygenic risk scores

GWAS results from Kunkle et al (Kunkle *et al*., 2019) were used to calculate an Alzheimer’s disease PRS for each individual. We choose this GWAS as it is based on clinically defined cases compared to controls. To separate the effects of *APOE* from other genetic variants associated with Alzheimer’s disease, we excluded the *APOE* region (chr19:45,116,911– 46,318,605) (Kunkle *et al*., 2019) from the PRS calculations. We generated PRS using PRSice (v2.0)(Choi and O’Reilly, 2019) which ‘clumps’ the Alzheimer’s disease GWAS summary statistics using the BDR genotype data such that the most significant variant in each LD block was retained. The PRS was then calculated in the target (BDR) dataset for each individual, as the number of reference alleles multiplied by the log odds ratio for that SNP (taken from the Kunkle et al Alzheimer’s disease GWAS), and then summed across all retained clumped variants with an Alzheimer’s disease GWAS P value < P_T._ A range of P value thresholds (P^T^) were used initially, to generate multiple possible PRS, where the optimal PRS was selected as the score that explained the highest proportion of variance (Nagelkerke’s pseudo R^2^) in Alzheimer’s disease case control status. In this analysis, Alzheimer’s disease cases and controls were defined as Braak high (Braak tangle stages V-VI) and low (Braak tangle stages 0-II) respectively, and PRS was tested using a logistic regression model with the first 8 genetic principal components as covariates. In the BDR, cohort the optimal threshold for selecting SNPs for the PRS was P < 5×10^™8^ (**Supplementary Fig. 2**). Prior to analysis the PRS calculated at this threshold was standardized to have a mean of 0 and SD of 1; therefore the interpretation is in units of SDs.

### APOE genotyping

The *APOE* SNPs rs7412 and rs429358 were genotyped with TaqMan assays using standard protocols. Where *APOE* genotype by TaqMan assay was not available, it was generated from the NeuroChip data (n = 44). The NeuroChip array includes multiple probes to assay the two *APOE* SNPs; based on the optimal concordance with the Taqman assay (91% concordant across assays) we used the probes rs7412.B3 and rs429358.T2 to determine *APOE* status. In all statistical analyses, *APOE* status was modelled as two numeric variables counting the number ε2 alleles and number of ε4 alleles an individual had. Given the rarity of ε2/ε2 genotype (only 4 occurrences (0.58%) in this sample), the ε2/ε2 individuals were combined with the individuals with one ε2 allele.

### Statistical analysis

All statistical analyses were performed in R version 3.5.2. All analytical code is available via GitHub (https://github.com/ejh243/BDR-Genetic-Analyses).

### Survival analysis

To test whether *APOE* and Alzheimer’s disease PRS were associated with younger age at death, we fitted Cox’s proportional hazards models using the R package survival. Three models were fitted with age at death as the outcome to test 1) *APOE* genotype modelled as two variables, 2) Alzheimer’s disease PRS and 3) *APOE* genotype and Alzheimer’s disease PRS simultaneously. All models included covariates for sex, BDR centre and 8 genetic principal components.

### Genetic analysis of neuropathology and clinical/cognitive status at death

Genetic associations between either *APOE* status or Alzheimer’s disease PRS and any of the continuous neuropathology variables (Braak tangle stage, Thal β-amyloid stage, CERAD stage, Braak Lewy body stage), clinical (CDR global rating) or cognitive status at death (MMSE, MoCA) were tested using a linear regression model. TDP-43 proteinopathy as a binary variable was analysed with logistic regression, but the model framework was the same. Up to four regression models were fitted for each variable. First, the effects of *APOE* status and Alzheimer’s disease PRS were estimated separately using Model 1 and Model 2 below.

Model 1:

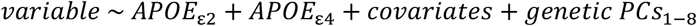

Model 2:

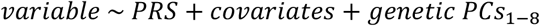

If *APOE* (either variable) and PRS were significantly associated with an outcome, then a multiple regression analysis was additionally fitted testing *APOE* and PRS simultaneously to confirm these were independent associations (Model 3).

Model 3:

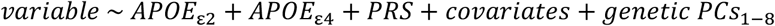

Finally, an interaction model (Model 4) between *APOE* and PRS was fitted to test if PRS associations differed depending on *APOE* genotype.

Model 4:

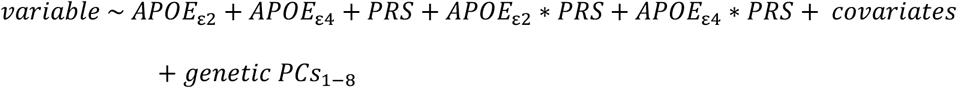

All analyses included age at death, sex, and BDR centre as covariates and the first eight genetic principal components. Analyses for clinical or cognition measures also included a covariate that measured the time lapse between the last assessment and death.

### Longitudinal clinical and cognition analyses

To test how *APOE* and Alzheimer’s disease PRS affected clinical status and cognitive trajectories, we fitted multi-level regression models using all available pre-mortem assessment data. A time variable was created which measured the number of days after the first visit that an assessment took place. Each cognitive variable was then tested as the dependent variable against this time variable included as a fixed effect along with covariates for age, sex, BDR centre and the first eight genetic principal components and a random effect for individual. To test for genetic effects on the cognitive trajectory, either *APOE* (coded as two variables) or Alzheimer’s disease PRS, was included in the model as a main effect and as an interaction with time. Models were fitted using the R packages lme4 and lmTest.

### Multiple testing

In total, we tested 12 outcomes against 3 genetic variables. Our outcomes comprised five neuropathological variables, one clinical variable at death, two cognitive measures, one longitudinal clinical, two longitudinal cognitive measures and a survival analysis of age at death. Against these 12 outcomes, we tested 3 genetic variables (Alzheimer’s disease PRS and two variables to model *APOE* genotype). Therefore, we performed a multiple testing correction for 36 tests, reporting significant associations as those with P < 0.0014. Given the correlations between the neuropathological, clinical and cognitive variables this is likely to be a conservative approach.

## Results

### Both tauopathy and β-amyloidosis are present at high frequencies in the BDR cohort

In order to profile the effects of both *APOE* genotype and Alzheimer’s disease PRS, our analyses were limited to BDR donors who had undergone neuropathological assessment and had NeuroChip array data (n = 693, **Table 1**). The participants had a mean age at death of 83.5 years (SD = 9.34 years) and 52.8% were male. Consistent with epidemiological reports, females were significantly older at death than males (mean difference = 3.84 years; P = 4.87 × 10^™8^). Our genetic analyses focused on four semi-quantitative and one indicator neuropathology variable. In 672 samples neurofibrillary tangle (NFT) pathology was quantified using Braak NFT stage (Braak and Braak, 1991; Braak *et al*., 2006) with a mean of 3.76 (SD = 1.90). Two variables reflecting the extent of β-amyloidosis were considered: β-amyloid distribution was measured by Thal β-amyloid phase (Thal *et al*., 2002) with a mean value of 3.14 (SD = 1.78) across 612 individuals and neuritic plaque density was scored using the CERAD classification (Mirra *et al*., 1991; Montine *et al*., 2012) with a mean value of 1.72 (SD = 1.26) across 634 individuals. α-Synuclein pathology was quantified using Braak Lewy body stage, where across 634 individuals the mean was 1.36 (SD = 2.26). TDP-43 status was available for 658 individuals, with 150 (22.8%) individuals classed as being TDP-43 positive.

**Table 1.**
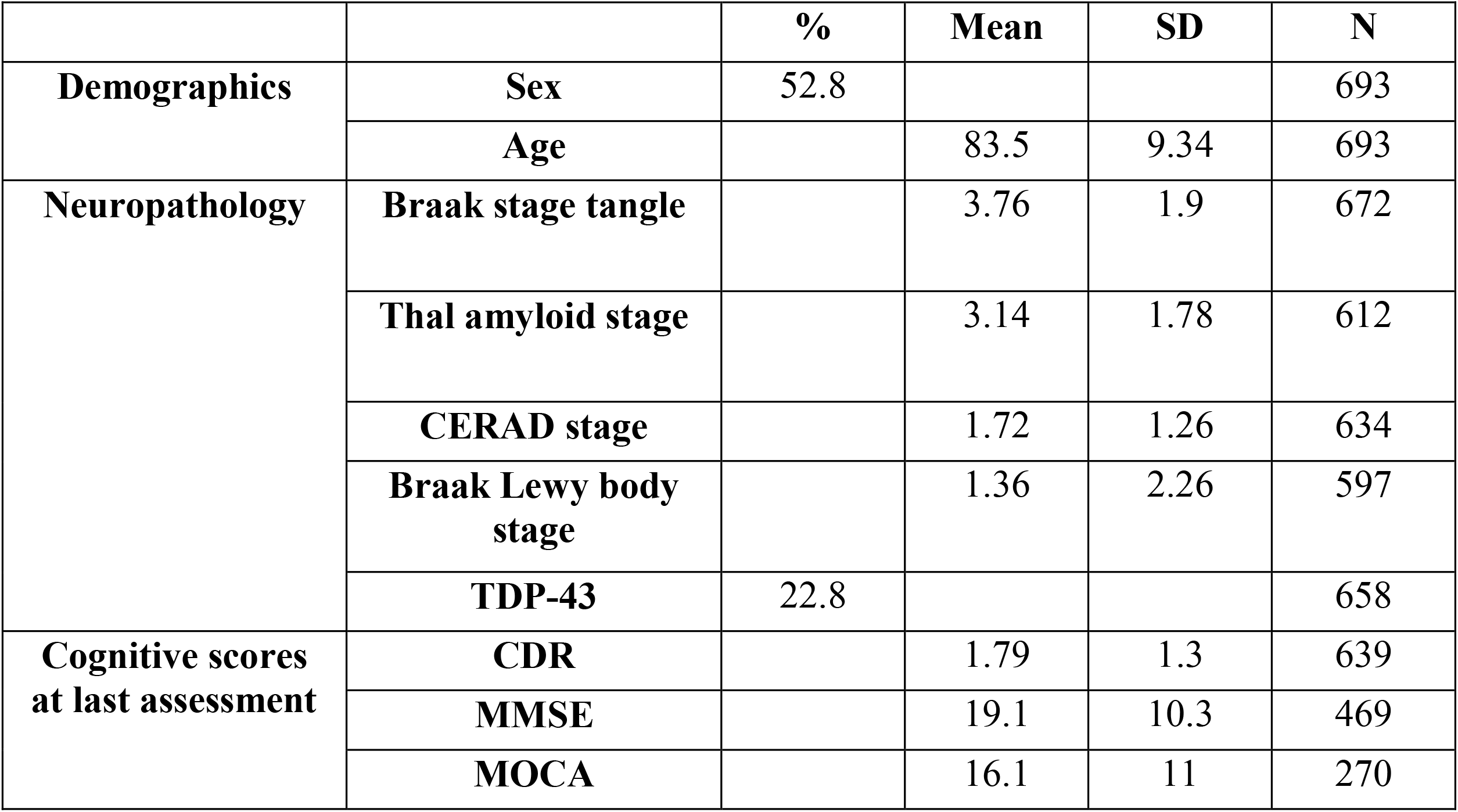
Summary of BDR cohort.

|  |  | % | Mean | SD | N |
| --- | --- | --- | --- | --- | --- |
| **Demographics** | **Sex** | 52.8 |  |  | 693 |
|  | **Age** |  | 83.5 | 9.34 | 693 |
| **Neuropathology** | **Braak stage tangle** |  | 3.76 | 1.9 | 672 |
|  | **Thal amyloid stage** |  | 3.14 | 1.78 | 612 |
|  | **CERAD stage** |  | 1.72 | 1.26 | 634 |
|  | **Braak Lewy body stage** |  | 1.36 | 2.26 | 597 |
|  | **TDP-43** | 22.8 |  |  | 658 |
| **Cognitive scores at last assessment** | **CDR** |  | 1.79 | 1.3 | 639 |
|  | **MMSE** |  | 19.1 | 10.3 | 469 |
|  | **MOCA** |  | 16.1 | 11 | 270 |

### Genetic risk factors for Alzheimer’s disease are associated with increased mortality

To determine whether higher genetic risk for Alzheimer’s disease was associated with increased mortality we analysed survival with Cox’s proportional hazard models (**Table 2**). *APOE* genotype was modelled as two variables – the number of ε4 alleles and the number of ε2 alleles, to distinguish the hypothesized risk effects of ε4 (Corder *et al*., 1993; Farrer *et al*., 1997) from the protective effects of ε2 (Reiman *et al*., 2020). Analysis of *APOE* genetic risk found that *APOE* ε4 status was significantly associated with younger age at death, with each additional ε4 allele associated with 29% increased risk of death (hazard ratio = 1.29; P = 9.66×10^™5^). Alzheimer’s disease PRS was nominally associated with an increased mortality (hazard ratio = 1.11; P = 8.97×10^™3^), although this was not significant after correcting for multiple testing.

**Table 2.** *APOE* is associated with increased mortality.

| Analytical model | APOE |  |  |  |  |  | Polygenic risk score |  |  |
| --- | --- | --- | --- | --- | --- | --- | --- | --- | --- |
|  | Number of ε2 alleles |  |  | Number of ε4 alleles |  |  |  |  |  |
|  | Hazard ratio | SE | P-value | Hazard ratio | SE | P-value | Hazard ratio | SE | P-value |
| Model 1 | 0.835 | 0.123 | 0.142 | 1.293 | 0.066 | 9.66E-05 |  |  |  |
| Model 2 |  |  |  |  |  |  | 1.105 | 0.038 | 8.97E-03 |
| Model 3 | 0.839 | 0.124 | 0.155 | 1.292 | 0.066 | 1.00E-04 | 1.106 | 0.038 | 8.41E-03 |

### APOE and Alzheimer’s disease PRS independently influence tauopathy and β-amyloidosis

The number of *APOE* ε4 alleles was positively associated (P < 0.00014) with all four semi-quantitative neuropathology measures (**Table 3**). The most significant association was with Braak neurofibrillary tangle (NFT) stage: each ε4 allele was associated with an increase of 1.16 Braak NFT stages (P = 4.16×10^™24^). Associations were also found between ε4 status and Thal β-amyloid phase (mean difference per ε4 allele = 0.981 phases; P = 3.96×10^™20^), neuritic plaque density (mean difference per ε4 allele = 0.713 stages; P = 1.03×10^™19^) and Braak Lewy body stage (mean difference per ε4 allele = 0.555 stages; P = 2.64×10^™4^). Alzheimer’s disease PRS was associated with two measures of neuropathology (**Table 3**): a higher polygenic burden was associated with Braak NFT stage (mean difference per SD of PRS = 0.354 stages; P = 1.36×10^™ 6^) and neuritic plaque density (mean difference per SD of PRS = 0.202 stages; P = 5.27×10^™5^). TDP-43 was not associated with either *APOE* genotype or Alzheimer’s disease PRS. Although variants in the *APOE* region were excluded from the PRS, we tested both *APOE* and PRS against Braak NFT stage and neuritic plaque density simultaneously to confirm that the identified associations were independent. The estimated effects of ε4 on both Braak NFT stage and neuritic plaque density were unaffected, while the Alzheimer’s disease PRS associations were slightly attenuated (**Table 3**) but remained significant. In addition to an additive model, we tested whether there was evidence for a multiplicative effect between Alzheimer’s disease PRS and *APOE* genotype on neuropathological burden to explore the hypothesis that in individuals with protective *APOE* genotypes, Alzheimer’s disease PRS is more important (i.e. has a larger effect on neuropathology). In this analysis, none of the five neuropathological variables had statistically significant differences across *APOE* genotype groups (P > 0.05) (**Supplementary Table 1**). Taken together, these results suggest that *APOE* status and Alzheimer’s disease PRS are independently associated with neuropathology, combining in an additive manner to influence an individual’s accumulation of tauopathy (NFTs) and β-amyloid plaques.

**Table 3.**
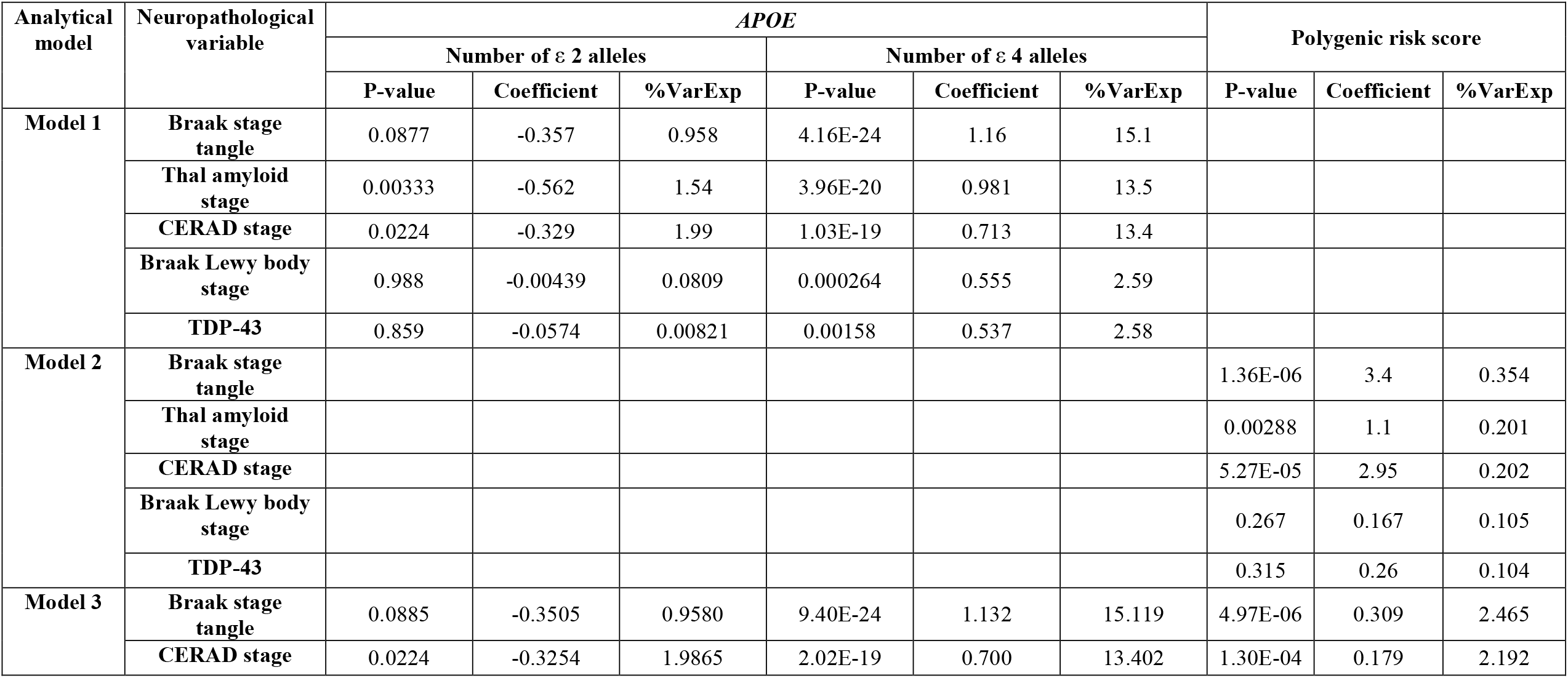
Common genetic risk factors for Alzheimer’s disease are associated with multiple aspects of neuropathology.

| Analytical model | Neuropathological variable | APOE |  |  |  |  |  | Polygenic risk score |  |  |
| --- | --- | --- | --- | --- | --- | --- | --- | --- | --- | --- |
|  |  | Number of ε 2 alleles |  |  | Number of ε 4 alleles |  |  |  |  |  |
|  |  | P-value | Coefficient | %VarExp | P-value | Coefficient | %VarExp | P-value | Coefficient | %VarExp |
| Model 1 | Braak stage tangle | 0.0877 | -0.357 | 0.958 | 4.16E-24 | 1.16 | 15.1 |  |  |  |
|  | Thal amyloid stage | 0.00333 | -0.562 | 1.54 | 3.96E-20 | 0.981 | 13.5 |  |  |  |
|  | CERAD stage | 0.0224 | -0.329 | 1.99 | 1.03E-19 | 0.713 | 13.4 |  |  |  |
|  | Braak Lewy body stage | 0.988 | -0.00439 | 0.0809 | 0.000264 | 0.555 | 2.59 |  |  |  |
|  | TDP-43 | 0.859 | -0.0574 | 0.00821 | 0.00158 | 0.537 | 2.58 |  |  |  |
| Model 2 | Braak stage tangle |  |  |  |  |  |  | 1.36E-06 | 3.4 | 0.354 |
|  | Thal amyloid stage |  |  |  |  |  |  | 0.00288 | 1.1 | 0.201 |
|  | CERAD stage |  |  |  |  |  |  | 5.27E-05 | 2.95 | 0.202 |
|  | Braak Lewy body stage |  |  |  |  |  |  | 0.267 | 0.167 | 0.105 |
|  | TDP-43 |  |  |  |  |  |  | 0.315 | 0.26 | 0.104 |
| Model 3 | Braak stage tangle | 0.0885 | -0.3505 | 0.9580 | 9.40E-24 | 1.132 | 15.119 | 4.97E-06 | 0.309 | 2.465 |
|  | CERAD stage | 0.0224 | -0.3254 | 1.9865 | 2.02E-19 | 0.700 | 13.402 | 1.30E-04 | 0.179 | 2.192 |

Given that the two distinct molecular pathologies - tauopathy and β-amyloidosis - that define Alzheimer’s disease are highly correlated (**Supplementary Fig. 3**), we wanted to establish whether *APOE* or Alzheimer’s disease PRS had a specific (or primary) effect on a particular aspect of neuropathology. To this end, we repeated the analysis of how Alzheimer’s disease PRS and *APOE* influence pathology, sequentially controlling for other neuropathology variables. This analysis revealed some interesting patterns. First, after controlling for any of the other three quantitative neuropathological variables, Braak Lewy body stage was not significantly associated with *APOE* ε4 (**Supplementary Table 2**) suggesting that the association we detected was largely driven by the fact that individuals with Lewy bodies also have NFTs and β-amyloid plaques. Second, after we controlled for Braak NFT stage, neither of the plaque measures remained significantly associated with *APOE* ε4 (**Supplementary Table 2**). In contrast, Braak NFT stage remained significantly associated with *APOE* ε4 status after controlling for plaque variable (adjusted for Thal phase, mean difference per *APOE* ε4 allele = 0.468; P = 6.44×10^™7^; adjusted for neuritic plaque density, mean difference per ε4 allele = 0.238; P = 1.82×10^™4^), albeit with an attenuated magnitude of effect. Considering the two measures of plaque burden, only Thal β-amyloid phase remained significantly associated with ε4 after controlling for neuritic plaque density (mean difference per ε4 allele = 0.265; P = 3.42×10^™4^). Neither Braak NFT stage nor neuritic plaque density remained significantly associated with Alzheimer’s disease PRS after controlling for the other measure of pathology (**Supplementary Table 2**). These results indicate that *APOE* ε4 has a specific influence on tauopathy (NFTs) as well as a shared effect on both plaque and NFT development, whereas the PRS is more generally associated with an increased burden of Alzheimer’s disease neuropathology.

### Association between APOE and cognitive performance is confounded by neuropathology

We determined clinical and cognitive status at death from the final pre-mortem assessment (**Table 1**). Data were available from 639 individuals who had had at least one CDR assessment with a mean final score of 1.79 (SD = 1.30) measured a mean of 353 days (SD = 374 days) prior to death. In addition, 469 individuals had had at least one MMSE assessment with a mean final score of 19.1 (SD = 10.3) measured a mean of 594 days (SD = 521 days) prior to death and 270 individuals had a MoCA assessment (mean = 16.1; SD = 11.0) measured a mean of 617 days (SD = 590 days) prior to death. *APOE* was significantly associated with dementia severity with each ε4 allele associated with an increase of 0.492 (P = 2.14×10^™9^) in pre-mortem CDR score (**Table 4**). *APOE* was also significantly associated with lower cognitive performance in MMSE prior to death (**Table 4**) with each ε4 allele being associated with a decrease of 4.86 (P = 1.30×10^™8^). In contrast, Alzheimer’s disease PRS was not significantly associated with any of the measures of clinical or cognitive status prior to death. To test whether the association between *APOE* and clinical measures was mediated by neuropathology we repeated these analyses including Braak NFT stage as an additional covariate; this variable had the largest effect in the genetic analyses described above, and its effect additionally captured associations with plaque pathology. In this model, the associations between *APOE* ε4 and CDR or MMSE were attenuated and neither remained significant (**Supplementary Table 3**). In contrast, on retesting Braak NFT stage whilst controlling for the clinical variables in turn, we observed that *APOE* ε4 remained significantly associated (**Supplementary Table 3**). This indicates that the association between *APOE* and clinical variables is a consequence of an increased burden of neuropathology.

**Table 4.** *APOE* is associated with clinical and cognitive status at death.

| Analytical model | Cognitive variable | APOE |  |  |  |  |  | Polygenic risk score |  |  |
| --- | --- | --- | --- | --- | --- | --- | --- | --- | --- | --- |
|  |  | Number of ε 2 alleles |  |  | Number of ε 4 alleles |  |  |  |  |  |
|  |  | P-value | Coefficient | %VarExp | P-value | Coefficient | %VarExp | P-value | Coefficient | %VarExp |
| Model 1 | CDR | 0.706 | -0.058 | 0.336 | 2.14E-09 | 0.492 | 9.83 |  |  |  |
|  | MMSE | 0.693 | 0.574 | 0.310 | 1.30E-08 | -4.859 | 10.05 |  |  |  |
|  | MOCA | 0.876 | -0.299 | 0.157 | 5.00E-03 | -3.403 | 3.19 |  |  |  |
| Model 2 | CDR |  |  |  |  |  |  | 0.034 | 0.109 | 1.82 |
|  | MMSE |  |  |  |  |  |  | 0.025 | -1.136 | 2.03 |
|  | MOCA |  |  |  |  |  |  | 0.785 | 0.191 | 0.089 |

### APOE ε4 is associated with faster cognitive decline in old age, but this is driven by Alzheimer’s disease neuropathological burden

Participants had a mean of 2.85 (SD = 1.71 visits) clinical assessment visits spread over a mean of 3.40 years (SD = 2.00 years) with a mean time between visits of 1.42 years (SD = 0.67 years). Over the course of all participants’ involvement in the BDR study, there was an overall decline in clinical status and cognitive performance. On average the CDR increased by a mean of 0.139 per year (P = 2.02×10^™31^), while MMSE declined by a mean of 1.07 per year (P = 3.00×10^™29^). *APOE* genotype was associated with worse cognitive scores at the start of the study and faster rates of decline as the study progressed (**Table 5**). For every ε4 allele, MMSE score was 3.19 points lower (P = 4.92×10^™5^) at the start of the study, and individuals then accumulated an additional decrease of 0.803 in their score per allele per year (P = 1.58×10^™8^). In contrast, although *APOE* was associated with a higher CDR score at the start of the study (mean difference per ε4 allele = 0.468; P = 4.34×10^™8^), there was no significant difference in the change in clinical status related to *APOE* as the study progressed. There was no significant association with MoCA scores and *APOE* genotype. There was no significant association between Alzheimer’s disease PRS and longitudinal clinical or cognitive profiles or clinical or cognitive status at study entry. On repeating these analyses using the participant’s age rather than time in the study, we found no significant linear associations with either cognitive status at study entry or performance as the study progressed (**Table 5**).

**Table 5.** *APOE* ε4 is associated with steeper cognitive decline prior to death.

| Time variable | Cognitive variable | Time |  | *APOE* |  |  |  | Interaction (Time x *APOE*) |  |  |  |
| --- | --- | --- | --- | --- | --- | --- | --- | --- | --- | --- | --- |
| | | | | Number of $\epsilon 2$ alleles | | Number of $\epsilon 4$ alleles | | Time x $\epsilon 2$ | | Time x $\epsilon 4$ | |
|  |  | Coefficient | P-value | Coefficient | P-value | Coefficient | P-value | Coefficient | P-value | Coefficient | P-value |
| Time since study entry (days) | CDR | 2.78E-04 | 2.45E-08 | 0.075 | 0.642 | 0.468 | 4.34E-08 | -1.34E-04 | 0.121 | 1.28E-04 | 9.83E-03 |
|  | MMSE | -1.39E-03 | 7.50E-05 | 1.209 | 0.371 | -3.195 | 4.92E-05 | -3.73E-04 | 0.529 | -2.20E-03 | 1.58E-08 |
|  | MOCA | -1.33E-03 | 0.146 | -0.663 | 0.710 | -2.335 | 0.040 | 1.98E-03 | 0.151 | -3.18E-03 | 0.024 |
| Age (years) | CDR | 2.01E-03 | 0.797 | -0.058 | 0.965 | -0.868 | 0.216 | 4.99E-04 | 0.974 | 0.017 | 0.042 |
|  | MMSE | -0.258 | 2.20E-04 | 4.759 | 0.675 | 2.574 | 0.689 | -0.040 | 0.766 | -0.086 | 0.268 |
|  | MOCA | 0.011 | 0.904 | -4.510 | 0.783 | -5.281 | 0.616 | 5.21E-02 | 0.785 | 0.031 | 0.809 |

Given our previous observation that genetic associations with clinical status and cognition are mediated by neuropathology, we wanted to confirm whether the longitudinal analyses were similarly affected. First, we tested whether change in clinical status was associated with neuropathology measured by Braak NFT stage, independent of genetic status (**Supplementary Table 5**). As expected, those with higher levels of tangle pathology at death had a more severe clinical rating, even at the start of the study (mean difference in CDR per Braak NFT stage = 0.355; P = 7.30×10^™42^) and declined quicker; each additional Braak NFT stage was associated with an additional increase of 0.0247 in CDR per year (P = 3.99×10^™5^). We observed similar results for cognitive performance measured by MMSE; at study entry, each additional Braak NFT stage was associated with a decrease of 2.58 in MMSE score (P = 7.27×10^™26^) and participants accumulated an additional decrease of 0.384 in MMSE per Braak NFT stage per year (P = 3.90×10^™15^). Repeating the *APOE* analysis with a covariate for the potential confounder of neuropathology found that in line with the cross-sectional analyses, the associations with both clinical severity and cognition were no longer significant after adjusting for Braak NFT stage (**Supplementary Table 6**). These results suggest that cognitive performance prior to death, and even many years before death, is a consequence of accumulating Alzheimer’s disease neuropathology.

## Discussion

In this study, we used the longitudinal cognitive and neuropathological assessment data in the BDR cohort to investigate how genetic risk factors for Alzheimer’s disease influence the accumulation of β-amyloid plaques, tauopathy, synucleinopathy, and TDP-43 proteinopathy, and progressive decline in clinical status and cognitive performance. Our results indicate that *APOE* ε4 status has the most dramatic influence on tauopathy (NFT burden) and that although *APOE* genotype is also associated with β-amyloidosis, synucleinopathy and cognition, these relationships are largely confounded by their correlation with tangle burden. Furthermore, our results indicate that *APOE* has a specific direct effect on NFT independent of other neuropathologies. Although this finding contradicts the predictions of the ‘amyloid cascade hypothesis’ in which tau tangle formation is considered secondary to β-amyloid pathology (Hardy and Allsop, 1991; Selkoe, 1991), it is consistent with careful neuropathologic studies that show that tauopathy can precede beta-amyloidosis, at least in some brain areas (Duyckaerts, 2011). Our results also agree with previous research showing that although the influence of *APOE* on tau tangles is largely mediated indirectly through neurobiological pathways associated with β-amyloid, approximately one third of its influence on tangle development is via an alternative non-amyloid pathway (Yu *et al*., 2014). Our findings also support the 2-process model proposed by Mungas et al (Mungas *et al*., 2014), according to which neocortical NFTs are mediated by β-amyloid deposition and medial temporal lobe NFTs and may be the consequence of a separate age-associated process.

In our analysis of pathologies that frequently co-occur with the accumulation of β-amyloid and tauopathy, we replicated the positive association between Lewy body burden and the *APOE* ε4 allele (Tsuang *et al*., 2013; Beecham *et al*., 2014). However, when we adjusted for either β-amyloid or NFTs, this association was attenuated, indicating that in our sample, the association may be a consequence of the higher levels of tau and β-amyloid in individuals with Lewy bodies. It should be noted that the majority of participants in our study were free of any Lewy body pathology, with 423 individuals (70.8%) having a Braak Lewy body stage of 0. Therefore these analyses may be underpowered, particularly in the context of disentangling the effects on multiple correlated neuropathology variables. In addition, we were not able to replicate associations between *APOE* genotype and the presence of TDP-43 proteinopathy (Josephs *et al*., 2014; Yang *et al*., 2018), although the direction of effect was consistent with previous reports. Although TDP-43 proteinopathy was not infrequent in the BDR cohort, with 22.8% participants classed as positive, our simple binary classification may have decreased our power to detect an effect. Although BDR is not limited to a particular dementia subtype, and includes unaffected controls, Alzheimer’s disease is the most common form of dementia and therefore the sample is enriched for NFT and β-amyloid pathology. To truly establish whether *APOE* genotype has an independent, direct effect on the common comorbidities associated with Alzheimer’s disease, such as Lewy bodies and TDP-43 proteinopathy, we will likely require a larger number of samples to detect residual effects after accounting for correlations between neuropathological variables.

As well as our examination of associations with *APOE*, we tested the cumulative effect of common Alzheimer’s disease-associated genetic variants on neuropathology, clinical status and cognition. Given that individual variants only confer a small amount of additional risk, we used a combined PRS to improve power. In contrast to *APOE*, the Alzheimer’s disease PRS was associated with both independent and shared effects on tauopathy (NFTs) and β-amyloidosis perhaps suggesting that, in combination, common genetic variants have a broader, more general effect on the neuropathological burden present in Alzheimer’s disease. This contrasts with findings from a previous study testing the consequences of an Alzheimer’s disease PRS without *APOE*, which only reported a significant association with NFTs and not β-amyloid plaques (Felsky *et al*., 2018). Of note, in that study the PRS was based on an older GWAS with fewer significant association signals, and therefore our study might highlight the additional power derived using variants from the latest GWAS for Alzheimer’s disease. While leveraging multiple genetic variants into a single PRS is a powerful approach, particularly where sample sizes are small, it can be challenging to interpret shared associations. As the PRS is a harmonised variable generated in our case from seventeen genetic variants, our results could be explained by different subsets of variants being causally associated with the distinct pathologies. This explanation fits with results from previous studies that have tested individual SNPs associated with Alzheimer’s disease against multiple measures of neuropathology reporting some variants having specific effects, while others were associated with multiple aspects (Beecham *et al*., 2014; Mäkelä *et al*., 2018). Furthermore, it is likely that some genetic risk factors do not act via either plaques or tauopathy (NFTs), possibly affecting other aspects of neuropathology such as vascular disease which was not included in this study.

We found that clinical and cognitive status at study recruitment and prior to death, in addition to decline over the course of the study, are not directly associated with *APOE* genotype but are likely to be a consequence of neuropathological burden and in particular the accumulation of NFTs. This concurs with results from a previous study in a slightly larger cohort that focused specifically on episodic memory and non-episodic cognition (Yu *et al*., 2014). Alzheimer’s disease-associated cognitive decline is hypothesised to start as much as 17 years prior to death, with the rate of decline fastest in those with the most extensive neuropathology; tauopathy, β-amyloidosis, TDP-43 proteinopathy, and synucleinopathy are all positively associated with decline (Boyle *et al*., 2017). While a strength of our study is the availability of longitudinal cognitive data, clinical data was only available for up to three years before death, limiting our ability to characterise the effects of neuropathology on cognitive trajectories. Furthermore, multiple aspects of neuropathology have been independently negatively associated with cognitive performance (Boyle *et al*., 2013). Although Alzheimer’s disease is characterized by β-amyloidosis and tauopathy, it is increasingly apparent that in older cohorts, there may be additional comorbidities which potentially confound this relationship (Schneider *et al*., 2009; James *et al*., 2012; James *et al*., 2016; Robinson *et al*., 2018). At present, the presence of multiple comorbidities makes it difficult to resolve cause from effect as each comorbidity may affect different domains of cognition at different times during pathogenesis. When considering the regional presence and global burden of different pathologies, there is extensive variation in the specific combination of neuropathological features that an individual develops ultimately having a unique effect on their individual cognitive performance over time (Boyle *et al*., 2018). The strengths of the BDR study design, collating repeated measures of cognitive performance in addition to standardized protocols for high quality neuropathological assessments in a large sample size make it an ideal dataset to ultimately disentangle the role of mixed pathologies on cognition and dementia and more extensive analyses will be possible in the future.

Our results should be considered in light of a number of limitations. First, the participants were self-selecting, which in line with many other observational cohorts introduces bias into the sample; they are from less deprived socio-economic areas and have higher levels of education than the general population. Second, consistent with the majority of genetic studies, our analysis was limited to participants of European ancestry to remove the biases associated with population stratification. Third, we only included a subset of Alzheimer’s disease and related neuropathology phenotypes, which were selected for practical reasons in that they were observed with sufficient frequency in the current sample. Analyses of rarer phenotypes will be possible with subsequent waves of the data as the overall sample size and number of cases increases. Fourth, our measures were of global cognition, rather than specific domains. As previous studies have found that different pathologies have specific effects of different cognitive domains (Yu *et al*., 2014), this may mean we miss some of the nuances of the relationship between neuropathology and cognition. Fifth, to aid interpretation of the analytical models we converted semi-quantitative neuropathological variables into continuous variables which assume an equal effect between all pairs of consecutive stages. This simplification may obscure some more complex patterns in the data but should enable us to pick up general correlations which were our primary interest. Finally, we did not control for severity of ischaemic brain damage, which is common in Alzheimer’s disease cases and negatively influences cognition.

In summary, our data indicate that *APOE* influences Alzheimer’s disease neuropathology via two independent pathways, one where β-amyloid accumulation mediates the development of tauopathy (NFTs), and a second pathway with direct effects on NFTs independent of β-amyloidosis. It is as a consequence of these neuropathological changes that cognitive performance is then impaired. The relationship between common genetic variants associated with Alzheimer’s disease and neuropathology is more complex, with each individual variant potentially having a different effect on neuropathology and cognition. Taken together, these results provide insights into how the symptoms of Alzheimer’s disease dementia manifest and how genetic risk factors influence the development of pathology.

## Data Availability

Genetic, clinical and cognitive data are available through the Dementias Platform UK (DPUK; https://www.dementiasplatform.uk/) platform upon application.

## Data Availability

Genetic, clinical and cognitive data are available through the Dementia’s Platform UK (DPUK; https://www.dementiasplatform.uk/) platform upon application.

## Acknowledgements

We would like to gratefully acknowledge all donors and their families for the tissue provided for this study. Human post-mortem tissue was obtained from the South West Dementia Brain Bank, London Neurodegenerative Diseases Brain Bank, Manchester Brain Bank, Newcastle Brain Tissue Resource and Oxford Brain Bank, members of the Brains for Dementia Research (BDR) Network. We wish to acknowledge the neuropathologists at each centre and BDR Brain Bank staff for the collection and classification of the samples.

## Abbreviations

BDR: Brains for Dementia Research
CDR: Clinical Dementia Rating
CERAD: Consortium to Establish a Registry for Alzheimer’s disease
GWAS: genome-wide association studies
LD: linkage disequilibrium
LOAD: late onset Alzheimer’s disease
MMSE: Mini-Mental State Examination
MoCA: Montreal Cognitive Assessment
NIA-AA: National Institute on Aging and Alzheimer’s Association
NFT: neurofibrillary tangle
PRS: polygenic risk score
QC: quality control
SD: standard deviation
SNP: single nucleotide polymorphism
TDP-43: transactive response DNA-binding protein 43

